# Association of Pre-pregnancy Mental Health Disorders with Hypertensive Disorders of Pregnancy

**DOI:** 10.1101/2025.02.03.25321596

**Authors:** Collette N. Ncube, Minhyuk Choi, Ruby Barnard-Mayers, Scott C. Zimmerman, Shelli Vodovozov, M. Maria Glymour, Samantha E. Parker

## Abstract

**Background:** Hypertensive disorders of pregnancy (HDP) affect nearly 10% of pregnancies in the U.S. and are known to increase risk of severe maternal morbidity and later life cardiovascular disease and stroke. Mental health disorders are common among pregnant individuals and have been linked to high blood pressure and adverse pregnancy outcomes. However, there is limited research investigating the association between history of mental health conditions and HDP.

**Objectives:** We aimed to assess the association of pre-pregnancy mental health disorders with HDP.

**Methods:** We used the NIH *All of Us* Research Program, which began enrollment in May 2018 and linked participants’ electronic health records (EHRs). First pregnancies with conception dates between October 2015 and September 2021 were identified from EHRs. Risk of HDP was examined among pregnancies of at least 20 weeks of gestation. We assessed the association between a pre-pregnancy mental health diagnosis and the risk of HDP using log-binomial regression models, estimating risk ratios (RR) and 95% confidence intervals, adjusted for racial and ethnic group and maternal age. We examined early pregnancy loss as a secondary outcome among all identified pregnancies. Results were stratified by the timing of pregnancy start relative to enrollment in the *All of Us* cohort.

**Results:** Among 11,756 pregnancies that occurred prior to study enrollment, we observed a 20% increased risk of HDP associated with pre-pregnancy anxiety and depression (RR 1.20, 95% CI 1.05,1.36) and a 50% increased risk of early pregnancy loss associated with pre-pregnancy anxiety and depression (RR 1.49, 95% CI 1.36,1.64). Among 2,358 pregnancies occurring after enrollment, findings for HDP were generally similar, but associations with early pregnancy loss were attenuated, with imprecise estimates.

**Conclusions:** Prior mental health disorder diagnoses were associated with a modest increase in risk of HDP and early pregnancy loss.

**SYNOPSIS:** *Study question:* Do pre-pregnancy mental health disorders increase the risk of pregnancy loss or hypertensive disorders of pregnancy (HDP)?

*What’s already known:* Few studies have evaluated pre-pregnancy mental health disorders and pregnancy loss. Findings from prior studies of the association between mental health disorders and HDP are inconsistent, with mental health status often ascertained in early pregnancy thus masking the temporal relationship with HDP.

*What this study adds:* In this study, which leverages the *All of Us* Research Program cohort, we report an increased risk of HDP associated with pre-pregnancy mental health disorders. Additionally, we found an increased risk of early pregnancy loss associated with pre-pregnancy mental health disorders, suggesting our results may underestimate the risk associated with HDP having conditioned on later gestations.

## Background

Hypertensive disorders of pregnancy (HDP), such as gestational hypertension and preeclampsia, complicate nearly 10% of pregnancies in the United States and are increasing in prevalence.^1^ HDP increases the risk of maternal and neonatal complications, including severe maternal morbidity, stillbirth, and preterm delivery. HDP also has long term implications for maternal health, including increased risks of cardiovascular disease development and mortality.^2–4^

The pathways leading to the development of HDP are multifactorial, which makes identifying intervention points for reducing rates and complications of HDP difficult. Risk factor identification for HDP has largely focused on pre-pregnancy and early pregnancy physical health, as well as reproductive history. There has been little attention devoted to understanding the impact of maternal mental health on HDP. Mental health disorders are common among pregnant individuals and have been linked to high blood pressure and adverse pregnancy outcomes, potentially due to shared risk factors such as stress and behavioral changes associated with mental health conditions.^5^ Animal studies have found a positive association between stress and vascular function, yet in population-based studies there is a lack of consistent evidence of the effects of stress (prior to and during pregnancy) on vascular function and hypertension-related diseases, likely due to differences in the ascertainment of stressors.^6–9^ Some studies have found that mental health disorders (including depression, anxiety, and bipolar disorder) diagnosed during early pregnancy are associated with HDP, but very few studies have assessed pre-pregnancy diagnoses. ^10–12^ Other studies have found no association between pre-existing mental health disorders and gestational hypertension but found an association with chronic hypertension. ^13,14^

Understanding the impact of mental health disorders on HDP is often complicated by the lack of data on the timing of onset of these conditions, thereby resulting in unclear temporality. Frequently, mental health disorders and HDP status are ascertained during pregnancy which makes establishing which condition came first difficult. This is particularly applicable to early pregnancy outcomes, such as spontaneous abortion or pregnancy loss. Complicated pregnancies and pregnancy loss are known to cause distress and therefore would be expected to be associated with mental health disorders in cross-sectional data. Pre-existing mental health disorders might contribute to pregnancy loss prior to 20 weeks of gestation. Previous research has found that mental health diagnoses such as depression and anxiety may contribute to spontaneous abortion risk. As hypertensive disorders of pregnancy are diagnosed at 20 weeks of gestation (at the earliest), pregnancies with fetal losses before 20 weeks would not be screened and thus not at risk for HDP.^12–15^

Understanding the relationship between mental health disorder history and HDP is essential for clinical care guidelines that support both maternal mental and physical health during pregnancy. Because early pregnancy loss by definition precludes incident HDP, to fully understand the influence of pre-pregnancy mental health disorders we must consider both early pregnancy loss and HDP. The primary objective of this study was to examine the association of pre-pregnancy mental health disorder diagnosis with HDP, and secondly with early pregnancy loss in a large and diverse cohort with longitudinal electronic health record data.

## Methods

### Cohort selection

We used data from the National Institutes of Health *All of Us* Research Program, described in detail elsewhere.^15^ Briefly, ongoing enrollment into the *All of Us* Research Program began May 2018, intentionally recruiting adults (18 years and older) from groups underrepresented in biomedical research. At enrollment, participants consent to share electronic health record (EHR) data (which includes their medical history prior to enrollment and is routinely updated after enrollment), complete surveys on sociodemographic and lifestyle factors, provide biospecimens and agree to physical assessments. Almost 850,000 participants have consented, and at least 570,000 of them enrolled as of mid-January 2025 and are being prospectively followed. Our study population includes all enrollees with EHR data, including individuals who enrolled during the beta testing phase which began May 2017.^16^ Participant EHR and survey data were obtained from the controlled-tier dataset [C2022Q4R9 v7], accessed via the Researcher Workbench.

To identify pregnancy episodes from EHR data in *All of Us,* we employed two algorithms. The first algorithm used R code from Smith et al.^17^, which is an adaptation of an algorithm originally developed by Jones et al.^18^ to identify pregnancies in the National COVID Cohort Collaborative. Briefly, this algorithm, called the Hierarchy and Rule-based Pregnancy Episode Inference integrated with Pregnancy Progression Signatures (HIPPS) algorithm identifies pregnancy episodes using records of delivery outcomes (Hierarchy-based Inference of Pregnancy (HIP) component) and gestational week codes (Pregnancy Progression Signature) and predicts a start and end date of the pregnancy.

Given the novelty of using All of Us for the epidemiologic study of pregnancy outcomes, our team created a secondary algorithm that prioritized gestational dating accuracy over the identification of all possible pregnancies within the database, by ascertaining pregnancy events using Z3A codes which are used to reflect gestational age. The pregnancy start date was estimated by subtracting the recorded weeks of gestation from the date of the Z3A record. As a secondary approach, we conducted the analyses among pregnancies identified using this algorithm alone. (see **Supplemental Table 1**).

From the pregnancies identified using the algorithms, we restricted our cohort to pregnancy episodes with a predicted conception date on/after October 1, 2015 and before September 1, 2021, which corresponds with the nationwide transition from International Classification of Diseases (ICD) 9th revision to the ICD 10th revision. The restriction pertaining to the earliest predicted conception date ensures consistent outcome ascertainment over the study period. Additionally, we applied an upper bound of September 1, 2021 for the pregnancy start date as we used the data released on the *All of Us* Researcher Workbench from July 1, 2022 and this restriction ensures sufficient time for delivery outcome assessment for all pregnancies. Lastly, we only included an individual’s first pregnancy in the EHR (i.e. index pregnancy). Data on parity are not available, therefore we used this restriction to approximate pregnancies among nulliparous individuals, to reduce the likelihood that capture of pre-pregnancy mental health assessment included mental health disorders related to prior deliveries (e.g. postpartum depression).

### Exposure

The exposure of interest was pre-pregnancy mental health disorders. We considered the following mental health disorders: depression, anxiety, bipolar disorder, post-traumatic stress disorder, obsessive-compulsive disorder, psychosis, and ‘other’ conditions (e.g. mood, stress, and sleep disorders) based upon the Alliance for Innovation on Maternal Health (AIM) Perinatal Mental Health Conditions list.^19,20^ We also created a combined ‘anxiety or depression’ category given the frequent co-occurrence of these conditions.^21^ An individual was determined to have pre-pregnancy mental health disorders if they had any of the associated diagnostic codes (see **Supplemental Table 2**) in the medical record time stamped prior to the start of the index pregnancy.

### Outcomes

The primary outcome was HDP, defined as a diagnosis of gestational hypertension, preeclampsia, eclampsia, HELLP syndrome, superimposed preeclampsia, or unspecified maternal hypertension during pregnancy. These conditions were captured using ICD-10-CM codes from the EHR (see **Supplemental Table 3**). Collectively, these codes have been shown to capture HDP with high sensitivity and specificity.^22^ One study found that ICD –9/10 codes had 94% sensitivity and specificity for preeclampsia, 85% sensitivity and 91% specificity for gestational hypertension, and 97% sensitivity and 85% specificity for any HDP.^23^ Diagnoses of HDP are made at or after 20 weeks of gestation, therefore we defined pregnancies at risk for HDP as those with documentation of a gestational period of 20 weeks or more, regardless of pregnancy outcome.

The secondary outcome was early pregnancy loss, defined as a pregnancy less than 140 days (20 weeks) in duration, including elective abortion, ectopic pregnancy, spontaneous abortion, or those with an unknown outcome but less than 20 weeks of gestation.

See **Supplementary Table 1** and **Supplementary Figure 1** for the list of ICD-10 codes and a decision tree used to identify pregnancy episodes and classify them based on risk of experiencing the primary or secondary outcomes.

### Covariates

Potential confounders of the association between mental health disorder history and HDP were selected based upon our causal assumptions about the shared determinants of mental health and HDP (assumptions are represented in the directed acyclic graph in **Figure 1**). Specifically, we considered a minimal set of covariates: maternal age (years) at the predicted date of conception and racial and ethnic identity (non-Hispanic Asian, non-Hispanic Black, Hispanic, non-Hispanic White, other). Participants classified as Asian, Black, Hispanic and White self-identified with only that racial/ethnic group, while the “other” category included individuals who identified as Middle Eastern or North African, Native Hawaiian or other Pacific Islander, and multi-racial or multi-ethnic, or another unspecified group. Given that many mental health conditions are pre-existing well before pregnancy start, we did not control for additional covariates that may have been influenced by the mental health conditions. ^24^

**Figure 1.**
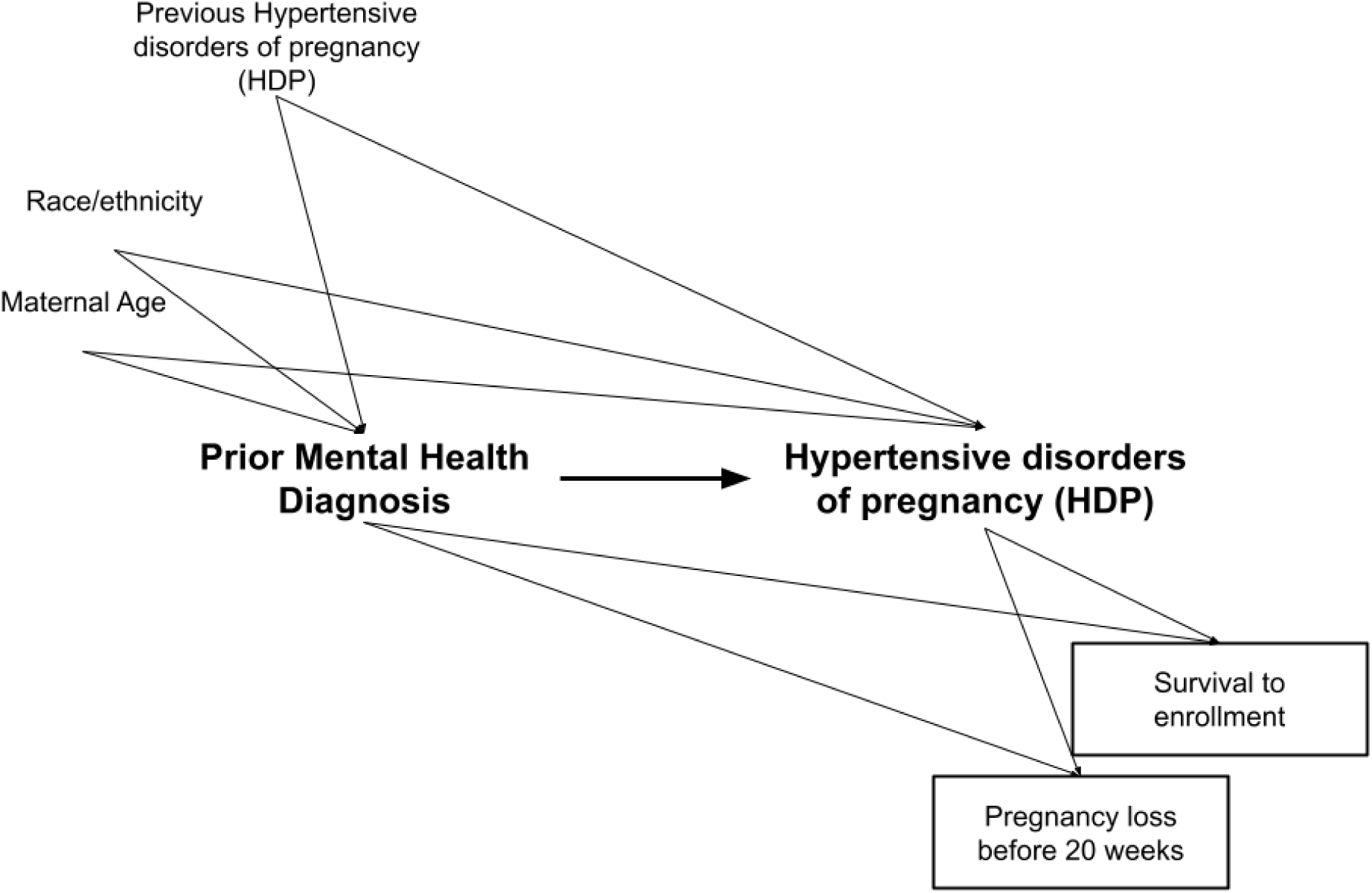
Directed Acyclic Graph Depicting the Relationship Between Mental Health Disorder Diagnosis and Hypertensive Disorders of Pregnancy.

### Statistical analysis

We described patient characteristics across pregnancy classifications: (1) overall, (2) pregnancies with HDP, (3) pregnancies of at least 20 weeks of gestation without HDP, and (4) pregnancies ending before 20 weeks of gestation. To assess both HDP and early pregnancy loss outcomes, we conducted log-binomial regression models to estimate risk ratios (RR) and 95% confidence intervals (CI). Among pregnancies that reached 20 weeks of gestation we estimated the risk of HDP comparing those with a diagnosed mental health condition to those without a diagnosis in the EHR prior to the predicted start of the index pregnancy. Among all pregnancies we estimated the risk of early pregnancy loss, comparing those with and without a recorded mental health disorder diagnoses prior to the predicted start of the index pregnancy. We stratified the analyses based upon the timing of the predicted start of the pregnancy in relation to enrollment in the *All of Us* research program (i.e., before or after enrollment), to address potential survival bias. All models were adjusted for maternal age at conception and racial and ethnic group.

In sensitivity analyses, we estimated the risk of HDP and early pregnancy loss among pregnancies identified and classified according to the secondary algorithm. All analyses were performed using R version 4.3.2. ^25^

### Missing data

The small amount of missing covariate data was handled using complete case analysis. Race and ethnicity information were missing for 1.9% (272/14,386) of pregnancies identified using the primary algorithm and for 1.6% (216/13,753) of pregnancies identified using the secondary algorithm. There were no missing values for maternal age. No significant differences were observed in the variables or outcomes before and after excluding the missing data (see **Supplemental Table 4**).

### Ethics approval

The *All of Us* Research Program is reviewed by an internal human subjects review board and all participants provide informed consent. Secondary analyses of de-identified data from the *All of Us* Research Program is not considered human subjects research.

## Results

From 413,457 participants, we identified 14,114 first pregnancy episodes using the primary algorithm and 13,753 first pregnancy episodes using the secondary algorithm with predicted conception dates between October 1, 2015 and September 1, 2021 (see **Figure 2**). Across the two algorithms we identified 11,223 pregnancy episodes in common, of which 10,631 (94.7%) were concordant regarding the classification of pregnancy outcome (see **Supplemental Table 5**).

**Figure 2.**
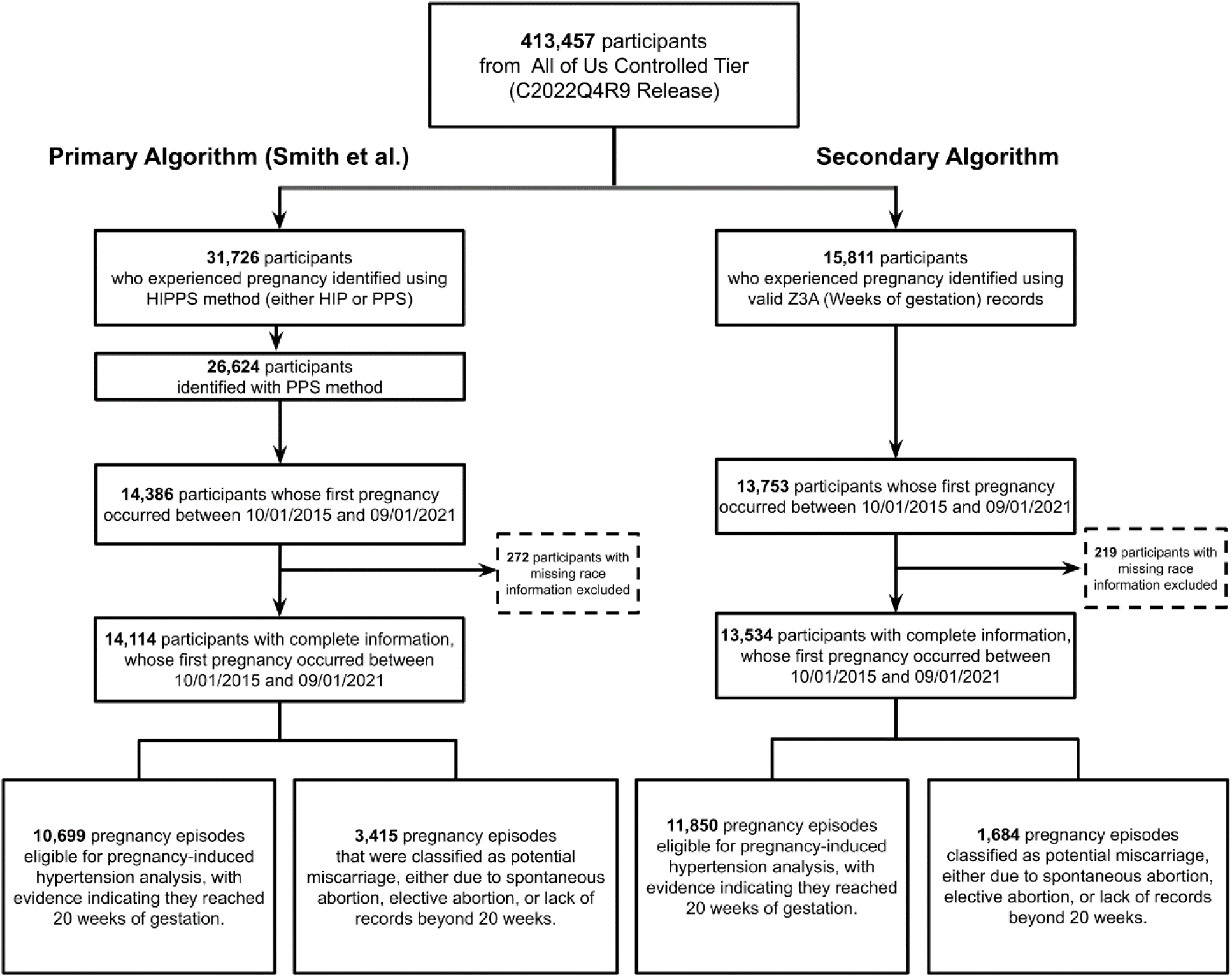
Flowchart of Participant Selection Based on Primary and Secondary Algorithms for Pregnancy Identification.

Among 14,114 pregnancy episodes identified in the primary algorithm (see **Table 1**), the mean (SD) age at pregnancy start was 30.1 (7.52) years, and 16.7% of identified pregnancy episodes occurred after *All of Us* enrollment. The racial and ethnic composition of the sample was as follows: 3.5% Asian, 15.0% Black, 39.9% Hispanic, 37.8% White, and 4.0% other. Almost a quarter of individuals with a pregnancy (22.0%) had a history of mental health disorders at pregnancy start, and 14.8% developed HDP. The characteristics of the 13,534 pregnancy episodes identified using the secondary algorithm were similar to those identified with the primary algorithm (see **Supplemental Table 6**).

**Table 1.** Participant Characteristics by Pregnancy Outcome Status, *All of Us* 2015-2022.

|  | **Overall**<br>(N=14,114) | **Hypertensive Disorders of Pregnancy<sup>a</sup>**<br>(N=2090) | **No Hypertensive Disorders of Pregnancy<sup>a</sup>**<br>(N=8609) | **Possible Early Pregnancy Loss<sup>b</sup>**<br>(N=3415) |
| --- | --- | --- | --- | --- |
| Maternal age (years), mean (SD) | 30.1 (7.52) | 29.1 (6.08) | 29.3 (6.89) | 33.0 (8.99) |
|  |  | *N (%)* |  |  |
| Racial and ethnic group |  |  |  |  |
| Asian | 489 (3.5) | 52 (2.5) | 330 (3.8) | 107 (3.1) |
| Black | 2111 (15.0) | 386 (18.5) | 1142 (13.3) | 583 (17.1) |
| Hispanic | 5626 (39.9) | 894 (42.8) | 3630 (42.2) | 1102 (32.3) |
| White | 5329 (37.8) | 679 (32.5) | 3167 (36.8) | 1483 (43.4) |
| Other | 559 (4.0) | 79 (3.8) | 340 (3.9) | 140 (4.1) |
| Pregnancy Timing |  |  |  |  |
| After enrollment | 2358 (16.7) | 318 (15.2) | 1117 (13.0) | 923 (27.0) |
| Before enrollment | 11756 (83.3) | 1772 (84.8) | 7492 (87.0) | 2492 (73.0) |
| Any Mental Health Disorder | 3106 (22.0) | 430 (20.6) | 1542 (17.9) | 1134 (33.2) |
| Anxiety or depression | 2826 (20.0) | 406 (19.4) | 1378 (16.0) | 1042 (30.5) |
| Anxiety | 2172 (15.4) | 320 (15.3) | 1033 (12.0) | 819 (24.0) |
| Depression | 1925 (13.6) | 267 (12.8) | 914 (10.6) | 744 (21.8) |
| Bipolar disorder | 564 (4.0) | 66 (3.2) | 264 (3.1) | 234 (6.9) |
| Obsessive compulsive disorder | 82 (0.6) | 13 (0.6) | 34 (0.4) | 35 (1.0) |
| Posttraumatic stress disorder | 355 (2.5) | 41 (2.0) | 154 (1.8) | 160 (4.7) |
| Psychosis | 192 (1.4) | 27 (1.3) | 81 (0.9) | 84 (2.5) |
| Other mental health disorder | 979 (6.9) | 117 (5.6) | 484 (5.6) | 378 (11.1) |

<sup>a</sup> Among pregnancies 20 weeks of gestation or more.
<sup>b</sup> Among pregnancies predicted to have ended prior to 20 weeks of gestation
*Note:* pregnancies were identified using the primary algorithm

Estimated associations between pre-pregnancy mental health disorders and HDP are presented in **Table 2** and **Figure 3**. Risk of HDP was elevated by approximately 20% if the mother had a pre-pregnancy diagnosis of anxiety or depression (RR 1.20, 95% CI: 1.05, 1.36), anxiety alone (RR 1.23, 95% CI: 1.07, 1.42), or depression alone (RR 1.19, 95% CI: 1.02, 1.38), compared to pregnancies in which the mother did not have each of these diagnoses. The estimates of HDP associated with the remaining mental health disorder types were imprecise, but either suggestive of an increased risk (for example, post-traumatic stress disorder RR 1.19, 95% CI: 0.80, 1.68), or null (for example, bipolar disorder RR 1.02, 95% CI: 0.76, 1.34). Among pregnancies that occurred after study enrollment, we observed similar, albeit imprecise estimates of the association between HDP and a diagnosis of anxiety or depression (RR 1.20, 95% CI 0.96, 1.51), anxiety alone (RR 1.31, 95% CI 1.03, 1.67), and depression alone (RR 1.16, 95% CI 0.88, 1.50)

**Figure 3.**
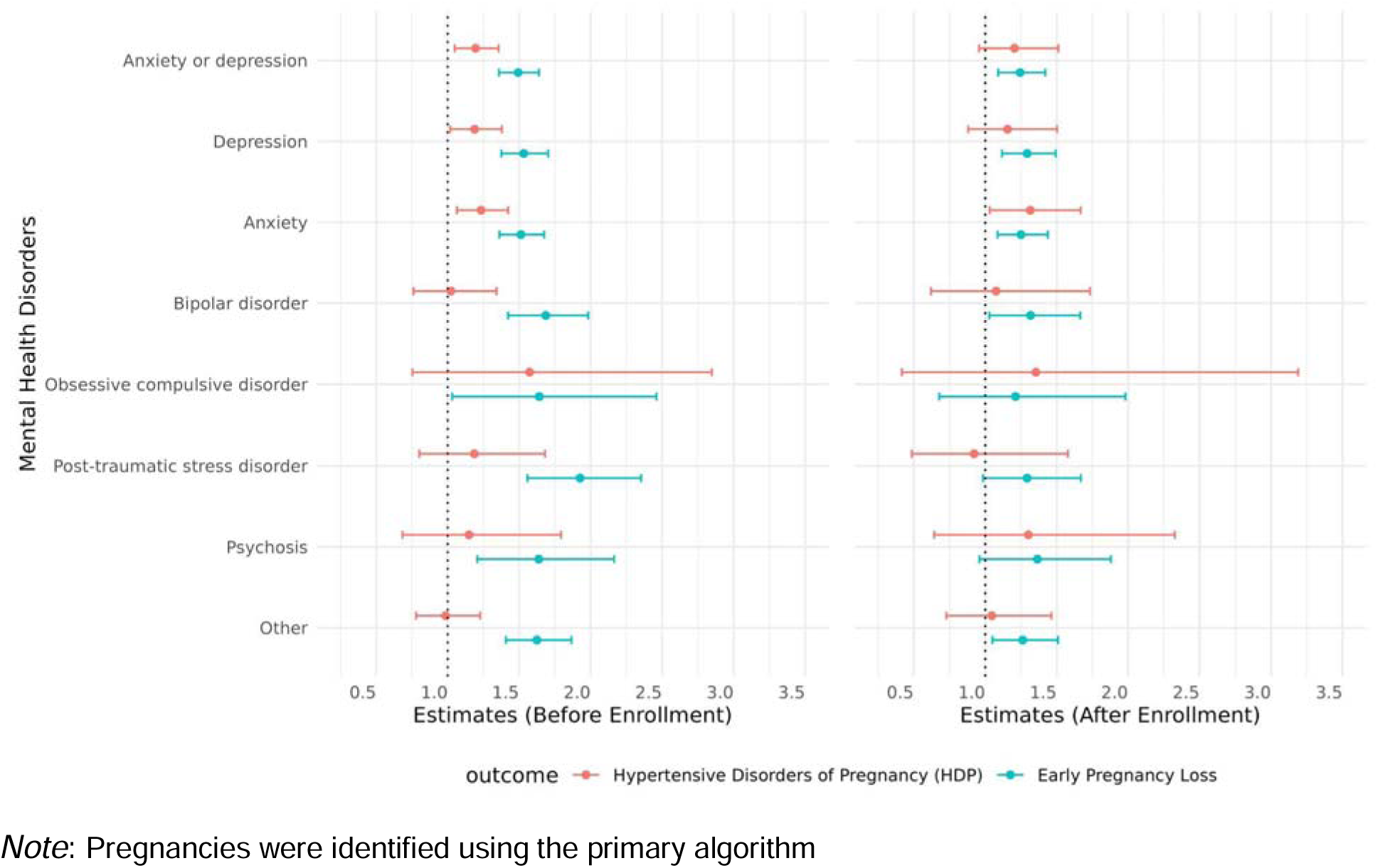
Association Between Diagnosed Mental Health Conditions and Hypertensive Disorders of Pregnancy or Early Pregnancy Loss, *All of Us* 2015-2022.

**Table 2.**
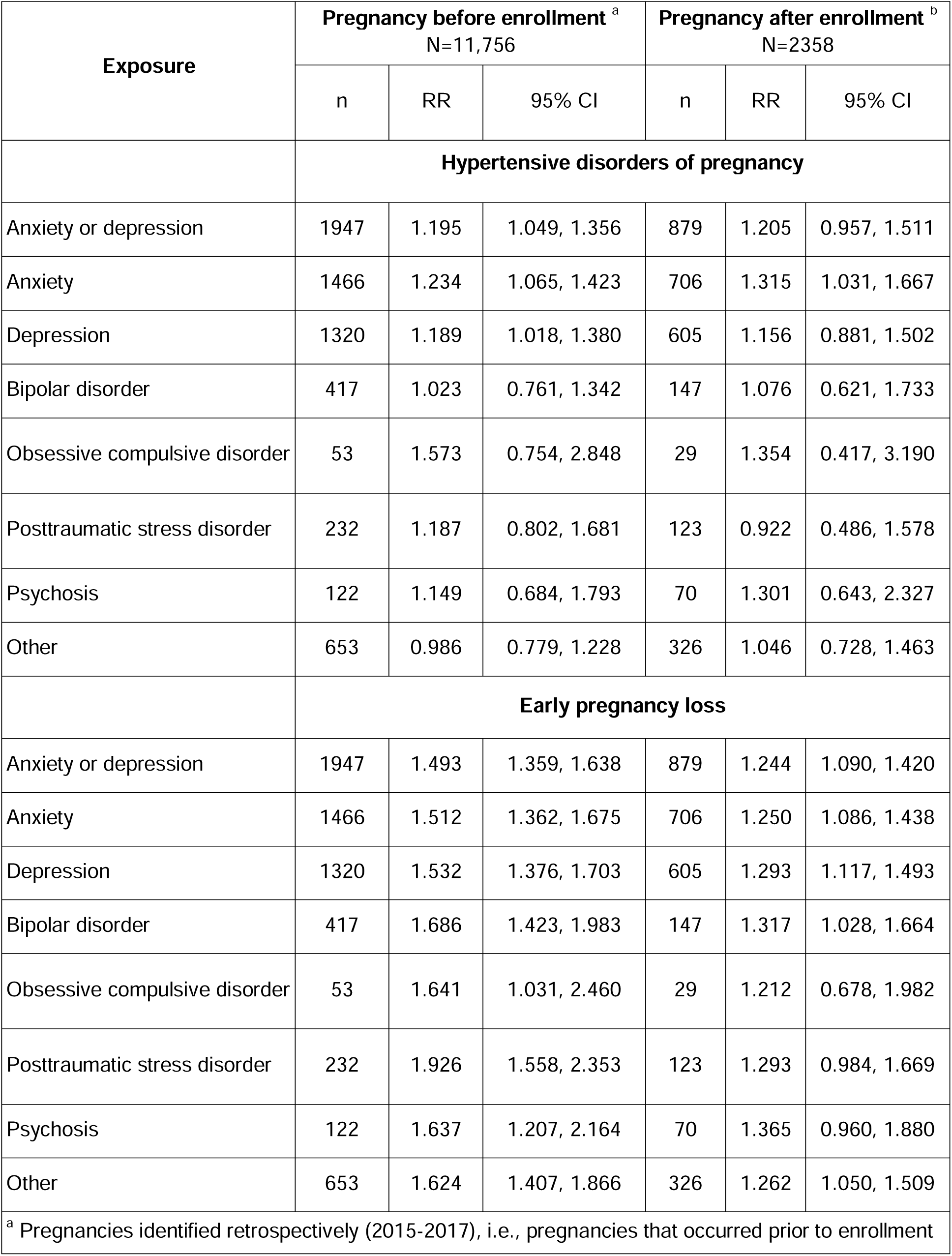
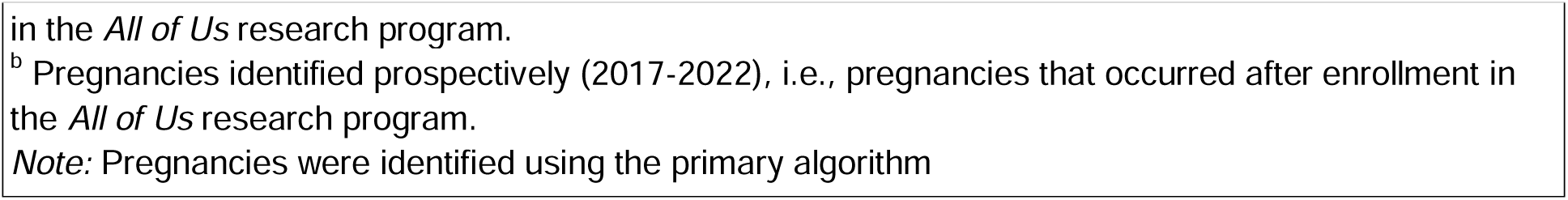
Association Between Diagnosed Mental Health Conditions and Hypertensive Disorders of Pregnancy or Early Pregnancy Loss, *All of Us* 2015-2022.

| Exposure | Pregnancy before enrollment <sup>a</sup><br>N=11,756 |  |  | Pregnancy after enrollment <sup>b</sup><br>N=2358 |  |  |
| --- | --- | --- | --- | --- | --- | --- |
|  | n | RR | 95% CI | n | RR | 95% CI |
|  | **Hypertensive disorders of pregnancy** |  |  |  |  |  |
| Anxiety or depression | 1947 | 1.195 | 1.049, 1.356 | 879 | 1.205 | 0.957, 1.511 |
| Anxiety | 1466 | 1.234 | 1.065, 1.423 | 706 | 1.315 | 1.031, 1.667 |
| Depression | 1320 | 1.189 | 1.018, 1.380 | 605 | 1.156 | 0.881, 1.502 |
| Bipolar disorder | 417 | 1.023 | 0.761, 1.342 | 147 | 1.076 | 0.621, 1.733 |
| Obsessive compulsive disorder | 53 | 1.573 | 0.754, 2.848 | 29 | 1.354 | 0.417, 3.190 |
| Posttraumatic stress disorder | 232 | 1.187 | 0.802, 1.681 | 123 | 0.922 | 0.486, 1.578 |
| Psychosis | 122 | 1.149 | 0.684, 1.793 | 70 | 1.301 | 0.643, 2.327 |
| Other | 653 | 0.986 | 0.779, 1.228 | 326 | 1.046 | 0.728, 1.463 |
|  | **Early pregnancy loss** |  |  |  |  |  |
| Anxiety or depression | 1947 | 1.493 | 1.359, 1.638 | 879 | 1.244 | 1.090, 1.420 |
| Anxiety | 1466 | 1.512 | 1.362, 1.675 | 706 | 1.250 | 1.086, 1.438 |
| Depression | 1320 | 1.532 | 1.376, 1.703 | 605 | 1.293 | 1.117, 1.493 |
| Bipolar disorder | 417 | 1.686 | 1.423, 1.983 | 147 | 1.317 | 1.028, 1.664 |
| Obsessive compulsive disorder | 53 | 1.641 | 1.031, 2.460 | 29 | 1.212 | 0.678, 1.982 |
| Posttraumatic stress disorder | 232 | 1.926 | 1.558, 2.353 | 123 | 1.293 | 0.984, 1.669 |
| Psychosis | 122 | 1.637 | 1.207, 2.164 | 70 | 1.365 | 0.960, 1.880 |
| Other | 653 | 1.624 | 1.407, 1.866 | 326 | 1.262 | 1.050, 1.509 |

<sup>a</sup> Pregnancies identified retrospectively (2015-2017), i.e., pregnancies that occurred prior to enrollment
in the *All of Us* research program.
<sup>b</sup> Pregnancies identified prospectively (2017-2022), i.e., pregnancies that occurred after enrollment in the *All of Us* research program.
*Note:* Pregnancies were identified using the primary algorithm

Among pregnancies that occurred prior to study enrollment, we observed at least a 50% increased risk of early pregnancy loss associated with all mental health disorder types (see **Table 2** and **Figure 3**), for example, a diagnosis of anxiety or depression (RR 1.49, 95% CI 1.36, 1.64), anxiety alone (RR 1.51, 95% CI 1.36 and 1.67), depression alone (RR 1.53, 95% CI 1.38, 1.70), and post-traumatic stress disorder (RR 1.93, 95% CI 1.56, 2.35). Among pregnancies that occurred after enrollment, the magnitudes of association were smaller but consistent with a 20% increased risk of early pregnancy loss, such as anxiety or depression (RR: 1.24, 95% CI: 1.09, 1.42) and bipolar disorder (RR 1.32, 95% CI 1.03, 1.66).

The results of sensitivity analyses using the secondary algorithm to identify pregnancy episodes are presented in **Supplementary Table 7** and **Supplementary Figure 2**. Overall, the findings were consistent with using the primary algorithm for pregnancy episode identification. Of note, the association between post-traumatic stress disorder and HDP was stronger among pregnancies that occurred prior to enrollment (RR 1.60, 95% CI: 1.20, 2.08). Other estimates associated with the risk of HDP and early pregnancy loss were similar within strata of pregnancies that occurred before and after enrollment.

### Comment

#### Principal findings

In a large and diverse cohort, select pre-pregnancy mental health disorders were associated with small increases in the risk of HDP. Depression and anxiety, the two most common mental health disorders, were associated with an approximate 20% increased risk of HDP. Risk ratios were also elevated for obsessive compulsive disorder and psychosis, but estimates were imprecise due to the small sample size. These findings were robust across several analyses, including examination of pregnancies documented in the EHR prior to and after enrollment in *All of Us* and use of two algorithms for the identification and gestational dating of pregnancies. The associations of pre-pregnancy mental health disorders with early pregnancy loss were even larger, with an almost a 50% increased risk of early pregnancy loss associated with pre-pregnancy depression and anxiety. The strong association between mental health disorder history and early pregnancy loss suggests that effects of mental health disorders on HDP may be underestimated due to the heightened pregnancy loss associated with these disorders. Although differences in point estimates associated with specific mental health disorder diagnoses were of a magnitude to be clinically relevant, the estimates were too imprecise to draw conclusions.

#### Strengths of the study

With the growing prevalence of mental health disorders, this evidence helps fill an important gap in research on the impact of maternal mental health on pregnancy outcomes. Prior studies have examined mental health disorders in relation to preterm delivery, stillbirth, and maternal morbidity, yet few studies have evaluated the association of pre-pregnancy mood and anxiety disorders with hypertension developed during pregnancy.^5^ Several studies have shown that mental disorders, depressive symptoms, or psychosocial stressors during pregnancy predict adverse outcomes, but these studies do not clearly establish temporal order.^12,26^ Our study specifically evaluated pre-pregnancy mental health and hypertensive disorders that onset during pregnancy, spotlighting the importance of women’s mental health across the lifecourse for shaping health changes during pregnancy.^27–29^ A single prior Canadian case control study evaluated an association between pre-pregnancy depression and HDP,^30^ finding results consistent with ours.

The ability to evaluate early pregnancy loss is an important strength of this study, augmenting interpretation of the results for HDP. The competing event of early pregnancy loss may lead to underestimations of the effect of pre-pregnancy mental health disorders on HDP. The strong association between pre-pregnancy mental health disorders and early pregnancy loss we documented is an important result and further strengthens the interpretation that pre-pregnancy mental health influences HDP.

Our sample is large and diverse, due to the *All of Us* Research Program’s enrollment efforts. A further strength of the present study is the use of EHR data, which includes dates of diagnoses, allowing for more accurate assessment of pre-pregnancy mental health conditions. We also used the AIM Perinatal Mental Health Condition ICD-10 codes for mental health conditions, with a crosswalk to ICD-9 codes in EHRs prior to October 1, 2015. Using this AIM list provides a standardized list of conditions and excludes pregnancy and postpartum specific diagnoses in an attempt to avoid capturing history of pregnancy-related transient mental health conditions.

An additional strength of our study is the comparison of two algorithms that approached pregnancy identification with different priorities. These algorithms were largely successful in identifying and dating the same pregnancy events, with some exceptions. The secondary algorithm did not capture many of the early pregnancy losses identified by the primary algorithm, while the primary algorithm did not capture many of the ongoing pregnancies at 20 weeks of gestation that were identified by the secondary algorithm. To better understand how the algorithms are sensitive to difference pregnancy outcomes, we also examined HDP and early pregnancy loss.

#### Limitations of the data

This study also has several limitations. Data on pregnancies are limited to those available in the EHRs shared with the *All of Us* Research Program, thereby missing data collected at healthcare encounters by providers either not EHRs or not providing their data for upload As such, we were unable to capture a comprehensive history of pregnancies, pregnancy outcomes, and complications among all participants. This also compromised our ability to distinguish with certainty pregnancies among nulliparous individuals from pregnancies among multiparous individuals for whom EHR records were not comprehensive. Parity data was not available in the EHR and not assessed in study surveys. Therefore, our selection of first reported pregnancies likely includes at least some participants who had given birth prior to the index pregnancy. Although some mental health diagnoses may have resulted from prior pregnancies (e.g. postpartum depression) and do not reflect chronic mental health conditions, this is unlikely to lead to substantial exposure misclassification. If engaged in care with an EHR-participating health care provider, a mental health disorder diagnosis is likely to be documented as psychiatric conditions often require ongoing treatment. The mean pre-pregnancy time with EHR data was 4.03 years; for individuals with a mental health disorder, an average of 20.29 (60.29) encounters with a diagnosis were documented. Other limitations of the analysis include lack of granular information on the mental health diagnosis, including severity and treatment. While we did examine the co-occurrence of depression and anxiety, we did not examine the impact of other co-morbid mental health conditions. Lastly, in our analysis of early pregnancy loss, we grouped outcomes of varying etiology including ectopic pregnancy, spontaneous abortion, and elective termination, therefore we are unable to comment on the role of mental health disorders on specific loss events, nor on losses occurring during the earliest gestational ages, as these are often undocumented in EHR data.

#### Interpretation

The results of our study are consistent with prior studies examining the association of pre-pregnancy depression or anxiety with hypertension in pregnancy. Thombre Kulkarni et al. reported small elevations in odds of incident HDP associated with anxiety and also depression, with wide CIs. Similarly, Serrano-Lomelin et al.^30^ reported that pre-pregnancy anxiety and depression were associated with slight elevations in gestational hypertension and pre-eclampsia. Few other studies have evaluated diagnosed mental health conditions prior to pregnancy and subsequent hypertension disorders developed in pregnancy, although several studies report on conditions during pregnancy or related constructs.^10,11^ Our findings, integrated with this prior evidence, strongly implicates mental health prior to pregnancy in shaping risk of hypertensive disorders developed during pregnancy. Anxiety and depression are both highly prevalent conditions in the population at large, suggesting treatment and management of mental health conditions are critical for promoting reproductive health.

Our findings regarding mental health diagnoses prior to pregnancy and pregnancy loss are also consistent with the limited prior research.^31^ Our primary focus in this paper was to interpret the effect of pre-pregnancy mental health disorders on HDP. To do so, it is essential to evaluate competing events, specifically early pregnancy loss, which by definition preclude developing HDP. Given that pre-pregnancy mental health disorders increase risk of early pregnancy loss, potential bias from this competing event likely underestimates the effects of pre-pregnancy mental health disorders on developing HDP.

## Conclusions

Key clinical implications of this research include care for pregnant people and care prior to pregnancy. Pregnant individuals with pre-pregnancy mental health conditions merit special attention for risk of pregnancy loss and complications related to hypertension. Currently, mental health conditions are not a recognized risk factor for aspirin prophylaxis for pre-eclampsia prevention. It may be beneficial to evaluate extending the indications for such preventive measures to include prior mental health conditions. Further, this research suggests that addressing the nationwide crisis of maternal health may entail addressing women’s health prior to pregnancy.

## Acknowledgments

The All of Us Research Program is supported by the National Institutes of Health, Office of the Director: Regional Medical Centers: 1 OT2 OD026549; 1 OT2 OD026554; 1 OT2 OD026557; 1 OT2 OD026556; 1 OT2 OD026550; 1 OT2 OD 026552; 1 OT2 OD026553; 1 OT2 OD026548; 1 OT2 OD026551; 1 OT2 OD026555; IAA #: AOD 16037; Federally Qualified Health Centers: HHSN 263201600085U; Data and Research Center: 5 U2C OD023196; Biobank: 1 U24 OD023121; The Participant Center: U24 OD023176; Participant Technology Systems Center: 1 U24 OD023163; Communications and Engagement: 3 OT2 OD023205; 3 OT2 OD023206; and Community Partners: 1 OT2 OD025277; 3 OT2 OD025315; 1 OT2 OD025337; 1 OT2 OD025276. In addition, the All of Us Research Program would not be possible without the partnership of its participants.

## Data Availability

This study utilized data from the Controlled Tier Dataset [v7] of the All of Us Research Program, accessible to authorized users on the Researcher Workbench. Statistical code used in our analyses are available from [GitHub repository link].

## Supplemental Files

**Supplemental Table 1:** Pregnancy Coding Categorization for Secondary Algorithm.

| Categorization | Codes |  |  |
| --- | --- | --- | --- |
|  | ICD-10 | ICD-10-PCS | CPT-4 |
| Category P1 (Clear Delivery) | Z37: Outcome of Delivery<br>Z38: Liveborn Infants According to Place of Birth and Type of Delivery<br>Z39: Encounter for Maternal Postpartum Care and Examination | 10E0XZZ: Delivery of Products of Conception, External Approach |  |
| Category P2 (Reached Delivery Stage but No Clear Outcome) | O47: False labor<br>O48: Late pregnancy<br>O60: Preterm labor<br>O61: Failed induction of labor<br>O62: Abnormalities of forces of labor<br>O63: Long labor<br>O64: Obstructed labor due to malposition and malpresentation of fetus<br>O65: Obstructed labor due to maternal pelvic abnormality<br>O66: Other obstructed labor<br>O67: Labor and delivery complicated by intrapartum hemorrhage<br>O68: Labor and delivery complicated by fetal stress<br>O69: Labor and delivery complicated by umbilical cord complications<br>O70: Perineal laceration during delivery<br>O71: Other obstetric trauma<br>O72: Postpartum hemorrhage<br>O73: Retained placenta and membranes, without hemorrhage<br>O74: Complications of anesthesia during labor and delivery<br>O75: Other complications of labor and delivery, not elsewhere classified<br>O80: Single spontaneous delivery<br>O81: Single delivery by forceps and vacuum extractor<br>O82: Single delivery by cesarean section<br>O85: Puerperal sepsis<br>O86: Other puerperal infections<br>O87: Venous complications in the puerperium<br>O88: Obstetric embolism<br>O89: Complications of anesthesia during the puerperium<br>O90: Complications of the puerperium, not elsewhere classified<br>O91: Infections of breast associated with childbirth<br>O92: Other disorders of breast and lactation associated with childbirth |  |  |
| Category P3 (Reached 20 Weeks) | Z3A.20~Z3A.49: weeks of gestations (20~49 weeks) |  |  |
| Category P4 (Elective Abortion) | O04:Complications following (induced) termination of pregnancy.<br>Z33.2:Encounter for elective termination of pregnancy | 10A07Z:Abortion | 01966: Anesthesia for procedures on the uterus, including induced abortion.<br>59850: Induced abortion by dilation and curettage (D&C).<br>59851: Induced abortion by dilation and evacuation (D&E).<br>59855: Induced abortion by intra-amniotic injection, including hospital admission and dilation and curettage or evacuation.<br>59856: Induced abortion by intra-amniotic injection, with hospital admission and termination of pregnancy.<br>59840: Induced abortion, dilation and curettage (D&C), outpatient setting.<br>59841: Induced abortion, dilation and evacuation (D&E), outpatient setting. |
| Category P5 (Early Pregnancy Loss or Ectopic pregnancy) | O00: Ectopic pregnancy<br>O02.1: Missed abortion<br>O20.0: Threatened abortion<br>O03: Spontaneous abortion<br>N96: Recurrent pregnancy loss |  |  |

**Supplemental Table 2.** ICD-10 codes for the diagnosis of perinatal mental health disorders.

| Diagnosis Codes |  |  |
| --- | --- | --- |
| Disorder type | ICD-10 codes | ICD-9 codes |
| Depression | F32: Depressive Episode (F32.0, F32.1, F32.2, F32.3, F32.4, F32.5, F32.8, F32.89, F32.9, F32.A)<br>F33: Major Depressive Disorder, Recurrent (F33.0, F33.1, F33.2, F33.3, F33.4, F33.40, F33.41, F33.42, F33.8, F33.9)<br>F34.1: Dysthymia | 293: Transient Mental Disorders Due to Conditions Classified Elsewhere (293.84)<br>300: Neurotic Disorders (300.00, 300.01, 300.02, 300.09, 300.20, 300.21, 300.22, 300.23, 300.29, 300.5, 300.89, 300.9)<br>306: Physiological Malfunctions Arising from Mental Factors (306.0, 306.1, 306.2, 306.3, 306.4, 306.50, 306.52, 306.53, 306.59, 306.7, 306.8)<br>308: Acute Reaction to Stress (308.9)<br>313: Disturbances of Emotion Specific to Childhood and Adolescence (313.0, 313.3, 313.82, 313.89)<br>V40: Mental and Behavioral Problems (V40.2, V40.9) |
| Bipolar disorder | F30: Manic Episode (F30.10, F30.11, F30.12, F30.13, F30.2, F30.3, F30.4, F30.8, F30.9)<br>F31: Bipolar Affective Disorder (F31.0, F31.10, F31.11, F31.12, F31.13, F31.2, F31.30, F31.31, F31.32, F31.4, F31.5, F31.60, F31.61, F31.62, F31.63, F31.64, F31.70, F31.71, F31.72, F31.73, F31.74, F31.75, F31.76, F31.77, F31.78, F31.81, F31.89, F31.9)<br>F34: Persistent Mood (Affective) Disorders (F34.0, F34.81, F34.89)<br>F39: Unspecified Mood (Affective) Disorder | 296: Episodic Mood Disorders (296.00, 296.01, 296.02, 296.03, 296.04, 296.05, 296.06, 296.40, 296.41, 296.42, 296.43, 296.44, 296.45, 296.46, 296.5, 296.51, 296.52, 296.53, 296.54, 296.55, 296.56, 296.60, 296.61, 296.62, 296.63, 296.64, 296.65, 296.66, 296.7, 296.80, 296.81, 296.89, 296.90, 296.99)<br>301.10: Affective personality disorder, unspecified<br>301.13: Cyclothymic disorder |
| Anxiety | F06.4: Organic Anxiety Disorder<br>F40: Phobic Anxiety Disorders (F40.00, F40.01, F40.02, F40.10, F40.11, F40.218, F40.240, F40.241, F40.8, F40.9)<br>F41: Other Anxiety Disorders (F41.0, F41.1, F41.3, F41.8, F41.9)<br>F43.0: Acute Stress Reaction<br>F45.8: Other Somatoform Disorders<br>F48: Other Neurotic Disorders (F48.8, F48.9)<br>F93.8: Other Childhood Emotional Disorders<br>F99: Mental Disorder, Not Otherwise Specified<br>R45.7: States of Emotional Shock and Stress, Unspecified | 296: Episodic Mood Disorders (296.20, 296.21, 296.22, 296.23, 296.24, 296.25, 296.26, 296.30, 296.31, 296.32, 296.33, 296.34, 296.35, 296.36, 296.82, 296.99)<br>298.0: Depressive type psychosis<br>300.4: Dysthymic disorder<br>301.12: Chronic hypomanic personality disorder<br>311: Depressive disorder, not elsewhere classified |
| Posttraumatic stress disorder | F43.1: Post-Traumatic Stress Disorder (PTSD) (F43.10, F43.11, F43.12) | 309.81: Post-traumatic stress disorder (PTSD) |
| Obsessive-compulsive disorder | F42: Obsessive-Compulsive Disorder (OCD) (F42.2, F42.3, F42.4, F42.8, F42.9)<br>R46.81: Obsessive-compulsive personality disorder | 300.3: Obsessive-compulsive disorder (OCD)<br>799.89: Other ill-defined conditions<br>V40.39: Other behavioral problems |
| Psychosis | F06: Other Mental Disorders Due to Known Physiological Conditions (F06.0, F06.2)<br>F20: Schizophrenia (F20.0, F20.1, F20.2, F20.3, F20.5, F20.81, F20.89, F20.9)<br>F21: Schizotypal Disorder<br>F22: Delusional Disorders<br>F23: Brief Psychotic Disorder<br>F24: Shared Psychotic Disorder<br>F25: Schizoaffective Disorders (F25.0, F25.1, F25.8, F25.9)<br>F28: Other Specified Schizophrenia Spectrum and Other Psychotic Disorders<br>F29: Unspecified Schizophrenia Spectrum and Other Psychotic Disorders<br>F44: Dissociative and Conversion Disorders (F44.0, F44.1, F44.2, F44.81, F44.89, F44.9)<br>F48: Other Nonpsychotic Mental Disorders (F48.1, F48.2) | 293: Transient Mental Disorders Due to Conditions Classified Elsewhere (293.81, 293.82)<br>295: Schizophrenic Disorders (295.10, 295.20, 295.30, 295.40, 295.60, 295.70, 295.80, 295.90)<br>297: Paranoid States (297.0, 297.1, 297.2, 297.3)<br>298: Other Nonorganic Psychoses (298.2, 298.3, 298.4, 298.8, 298.9)<br>300: Neurotic Disorders (300.12, 300.13, 300.14, 300.15, 300.16, 300.19, 300.6)<br>301.22: Schizotypal personality disorder<br>310.81: Pseudobulbar affect |
| Other mental health disorders | R45: Symptoms and Signs Involving Emotional State (R45.850, R45.851)<br>F06: Other Mental Disorders Due to Known Physiological Conditions (F06.1, F06.30, F06.31, F06.32, F06.33, F06.34)<br>F34.9: Persistent mood (affective) disorder, unspecified<br>F43: Reaction to Severe Stress, and Adjustment Disorders (F43.20, F43.21, F43.22, F43.23, F43.24, F43.25, F43.29, F43.81, F43.89, F43.9)<br>F44: Dissociative and Conversion Disorders (F44.4, F44.5, F44.6, F44.7)<br>F45: Somatoform Disorders (F45.0, F45.1, F45.20, F45.21, F45.22, F45.29, F45.41, F45.42, F45.8, F45.9)<br>F51: Sleep Disorders Not Due to a Substance or Known Physiological Condition (F51.01, F51.02, F51.03, F51.04, F51.05, F51.09, F51.11, F51.12, F51.13, F51.19, F51.8, F51.9)<br>F54: Psychological and Behavioral Factors Associated with Disorders or Diseases Classified Elsewhere<br>F59: Unspecified Behavioral Syndromes Associated with Physiological Disturbances and Physical Factors | 293: Transient Mental Disorders Due to Conditions Classified Elsewhere (293.83, 293.89)<br>294.8: Other persistent mental disorders due to conditions classified elsewhere<br>296.99: Other specified bipolar disorder<br>300: Neurotic Disorders (300.11, 300.81, 300.82, 300.7, 300.89)<br>306: Physiological Malfunctions Arising from Mental Factors (306.0, 306.1, 306.2, 306.3, 306.4, 306.50, 306.52, 306.53, 306.59, 306.7, 306.8, 306.9)<br>307: Special Symptoms or Syndromes, Not Elsewhere Classified (307.40, 307.41, 307.42, 307.44, 307.49, 307.80, 307.89)<br>309: Adjustment Disorders (309.0, 309.1, 309.24, 309.28, 309.29, 309.3, 309.4, 309.89, 309.9)<br>316: Psychic Factors Associated with Diseases Classified Elsewhere<br>327: Organic Sleep Disorders (327.02, 327.15, 327.19)<br>V62: Problems Related to Life Circumstances (V62.84, V62.85) |

**Supplemental Table 3.** ICD-10 codes for the diagnosis of hypertensive disorders of pregnancy.

| Diagnosis Codes |  |
| --- | --- |
| Disorder type | ICD-10 codes |
| Superimposed preeclampsia | O11: Pre-existing Hypertension with Superimposed Proteinuria (O11, O11.1, O11.2, O11.3, O11.4, O11.5, O11.9) |
| Gestational hypertension (without significant proteinuria) | O13: Gestational Hypertension without Significant Proteinuria (O13, O13.1, O13.2, O13.3, O13.4, O13.5, O13.9) |
| Preeclampsia, mild to moderate | O14.0: Mild to Moderate Pre-eclampsia (O14.00, O14.02, O14.03, O14.04, O14.05) |
| Preeclampsia, severe | O14.1: Severe Pre-eclampsia (O14.10, O14.12, O14.13, O14.14, O14.15) |
| Preeclampsia, unspecified | O14.9: Pre-eclampsia, Unspecified (O14.90, O14.92, O14.93, O14.94, O14.95) |
| Eclampsia | O15: Eclampsia (O15.0, O15.00, O15.02, O15.03, O15.1, O15.2, O15.9) |
| HELLP syndrome | O14.2: HELLP Syndrome (Hemolysis, Elevated Liver enzymes, Low Platelets) (O14.20, O14.22, O14.23, O14.24, O14.25) |
| Maternal hypertension, unspecified | O16: Unspecified Maternal Hypertension (O16, O16.1, O16.2, O16.3, O16.4, O16.5, O16.9) |

**Supplemental Table 4.** Comparison of Variables and Outcomes Before and After Excluding Missing Data: Complete Case Analysis.

|  |  | Primary Algorithm |  | Secondary Algorithm |  |
| --- | --- | --- | --- | --- | --- |
|  | Variable name | Incomplete Case<br>(N=14,386) | Complete Case<br>(N=14,114) | Incomplete Case<br>(N=13,753) | Complete Case<br>(N=13,534) |
|  |  |  | *N (%)*<sup>a</sup> |  |  |
| Outcome | Pregnancy ≥ 20 weeks gestation, no HDP | 8770 (61.0) | 8609 (61.0) | 9422 (68.5) | 9273 (68.5) |
|  | Pregnancy ≥ 20 weeks gestation, HDP | 2121 (14.7) | 2090 (14.8) | 2620 (19.1) | 2577 (19.0) |
|  | Pregnancy predicted < 20 weeks gestation | 3495 (24.3) | 3415 (24.2) | 1711 (12.4) | 1684 (12.4) |
| Covariates | Maternal age, Mean (SD) | 30.2 (7.54) | 30.1 (7.52) | 29.2 (6.10) | 29.2 (6.09) |
|  | Racial and ethnic group |  |  |  |  |
|  | Asian | 489 (3.4) | 489 (3.5) | 455 (3.3) | 455 (3.4) |
|  | Black | 2111 (14.7) | 2111 (15.0) | 2005 (14.6) | 2005 (14.8) |
|  | Hispanic | 5626 (39.1) | 5626 (39.9) | 6039 (43.9) | 6039 (44.6) |
|  | White | 5329 (37.0) | 5329 (37.8) | 4498 (32.7) | 4498 (33.2) |
|  | Other | 559 (3.9) | 559 (4.0) | 537 (3.9) | 537 (4.0) |
|  | Unknown | 272 (1.9) | 0 (0.0) | 219 (1.6) | 0 (0.0) |
|  | Pregnancy Timing |  |  |  |  |
|  | After enrollment | 2398 (16.7) | 2358 (16.7) | 1967 (14.3) | 1945 (14.4) |
|  | Before enrollment | 11988 (83.3) | 11756 (83.3) | 11786 (85.7) | 11589 (85.6) |
| Exposure | Any Mental Health Disorder | 3177 (22.1) | 3106 (22.0) | 3076 (22.4) | 3021 (22.3) |
|  | Anxiety or depression | 2891 (20.1) | 2826 (20.0) | 2772 (20.2) | 2722 (20.1) |
|  | Anxiety | 2223 (15.5) | 2172 (15.4) | 2043 (14.9) | 2005 (14.8) |
|  | Depression | 1968 (13.7) | 1925 (13.6) | 1906 (13.9) | 1875 (13.9) |
|  | Bipolar disorder | 576 (4.0) | 564 (4.0) | 519 (3.8) | 510 (3.8) |
|  | Obsessive compulsive disorder | 83 (0.6) | 82 (0.6) | 63 (0.5) | 63 (0.5) |
|  | Posttraumatic stress disorder | 365 (2.5) | 355 (2.5) | 299 (2.2) | 290 (2.1) |
|  | Psychosis | 200 (1.4) | 192 (1.4) | 169 (1.2) | 165 (1.2) |
|  | Other | 1004 (7.0) | 979 (6.9) | 1083 (7.9) | 1060 (7.8) |
| <sup>a</sup> Except where noted |  |  |  |  |  |

**Supplemental Table 5.** Cross-Classification of Pregnancy Outcomes Identified by Primary and Secondary Algorithms.

|  |  | Secondary Algorithm |  |  |  |
| --- | --- | --- | --- | --- | --- |
|  | Primary Algorithm /<br>Secondary Algorithm | 1:Hypertensive<br>Disorders of Pregnancy<br>(HDP), made it to 20<br>weeks | 2:no Hypertensive<br>Disorders of Pregnancy<br>(HDP), made it to 20<br>weeks | 3:Possible Early<br>Pregnancy Loss<br>(Pregnancy predicted as<br>less than 20 weeks) | 4:not captured in<br>Secondary Algorithm |
| **Primary**<br>**Algorithm** | 1:Hypertensive<br>Disorders of Pregnancy<br>(HDP), made it to 20<br>weeks | 2024 | 9 | 3 | 54 |
|  | 2:no Hypertensive<br>Disorders of Pregnancy<br>(HDP), made it to 20<br>weeks | 16 | 7320 | 63 | 1210 |
|  | 3:Possible Early<br>Pregnancy Loss<br>(Pregnancy predicted as<br>less than 20 weeks) | 91 | 410 | 1287 | 1627 |
|  | 4:not captured in<br>Primary Algorithm | 446 | 1534 | 331 | 0 |

**Supplemental Table 6.**
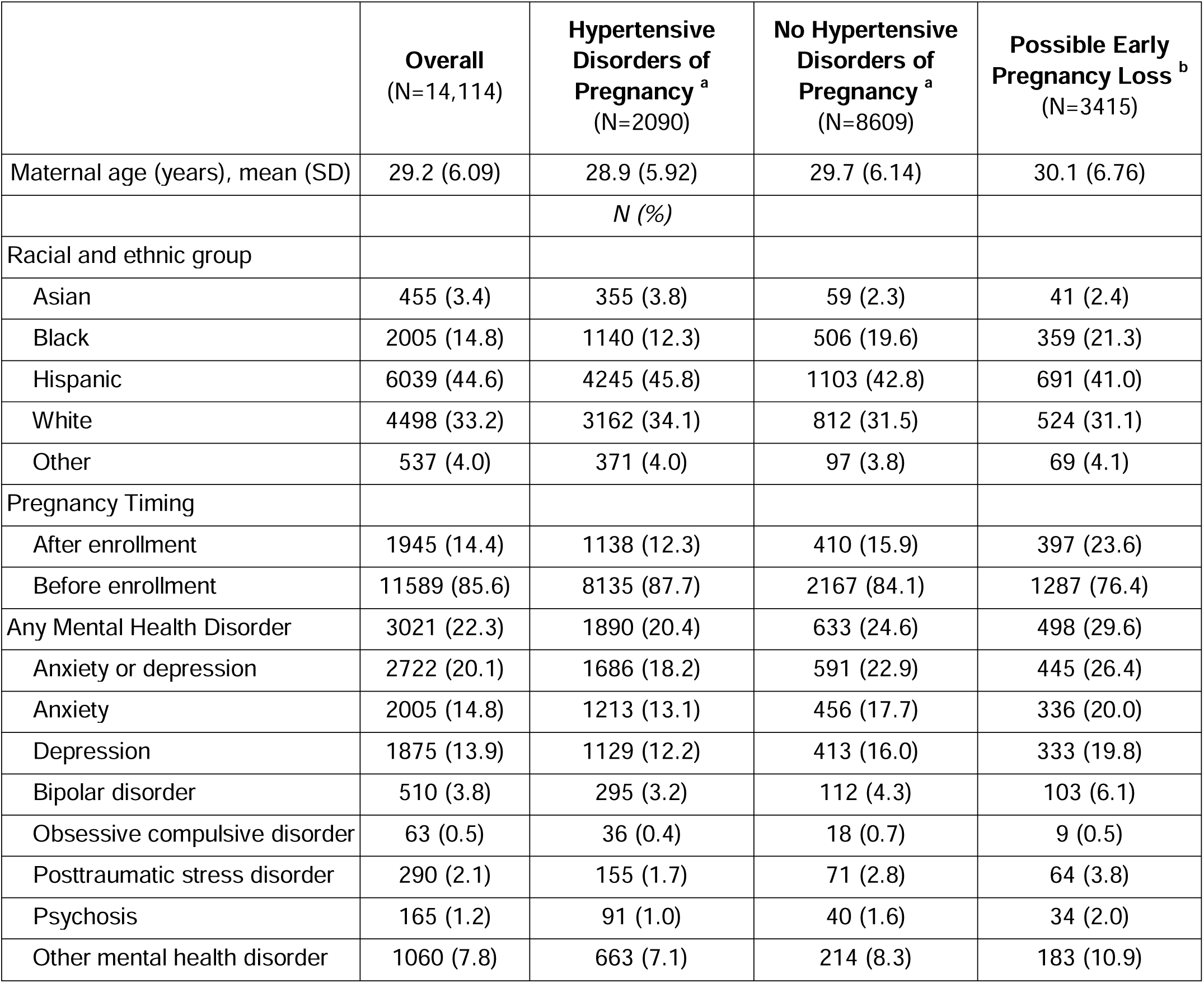
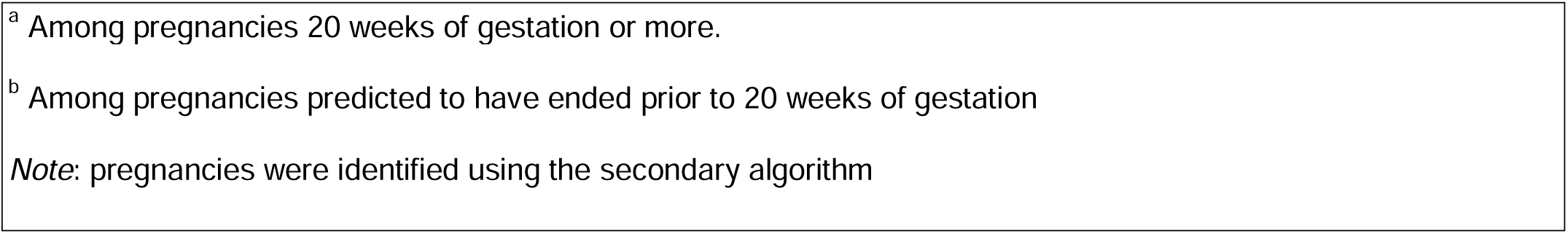
Characteristics of Participants by Pregnancy Outcome Status, Identified in the Secondary Algorithm, *All of Us* 2015-2022.

|  | **Overall**<br>(N=14,114) | **Hypertensive Disorders of Pregnancy <sup>a</sup>**<br>(N=2090) | **No Hypertensive Disorders of Pregnancy <sup>a</sup>**<br>(N=8609) | **Possible Early Pregnancy Loss <sup>b</sup>**<br>(N=3415) |
| --- | --- | --- | --- | --- |
| Maternal age (years), mean (SD) | 29.2 (6.09) | 28.9 (5.92) | 29.7 (6.14) | 30.1 (6.76) |
|  |  | *N (%)* |  |  |
| Racial and ethnic group |  |  |  |  |
| Asian | 455 (3.4) | 355 (3.8) | 59 (2.3) | 41 (2.4) |
| Black | 2005 (14.8) | 1140 (12.3) | 506 (19.6) | 359 (21.3) |
| Hispanic | 6039 (44.6) | 4245 (45.8) | 1103 (42.8) | 691 (41.0) |
| White | 4498 (33.2) | 3162 (34.1) | 812 (31.5) | 524 (31.1) |
| Other | 537 (4.0) | 371 (4.0) | 97 (3.8) | 69 (4.1) |
| Pregnancy Timing |  |  |  |  |
| After enrollment | 1945 (14.4) | 1138 (12.3) | 410 (15.9) | 397 (23.6) |
| Before enrollment | 11589 (85.6) | 8135 (87.7) | 2167 (84.1) | 1287 (76.4) |
| Any Mental Health Disorder | 3021 (22.3) | 1890 (20.4) | 633 (24.6) | 498 (29.6) |
| Anxiety or depression | 2722 (20.1) | 1686 (18.2) | 591 (22.9) | 445 (26.4) |
| Anxiety | 2005 (14.8) | 1213 (13.1) | 456 (17.7) | 336 (20.0) |
| Depression | 1875 (13.9) | 1129 (12.2) | 413 (16.0) | 333 (19.8) |
| Bipolar disorder | 510 (3.8) | 295 (3.2) | 112 (4.3) | 103 (6.1) |
| Obsessive compulsive disorder | 63 (0.5) | 36 (0.4) | 18 (0.7) | 9 (0.5) |
| Posttraumatic stress disorder | 290 (2.1) | 155 (1.7) | 71 (2.8) | 64 (3.8) |
| Psychosis | 165 (1.2) | 91 (1.0) | 40 (1.6) | 34 (2.0) |
| Other mental health disorder | 1060 (7.8) | 663 (7.1) | 214 (8.3) | 183 (10.9) |

<sup>a</sup> Among pregnancies 20 weeks of gestation or more.
<sup>b</sup> Among pregnancies predicted to have ended prior to 20 weeks of gestation
*Note:* pregnancies were identified using the secondary algorithm

**Supplemental Table 7.**
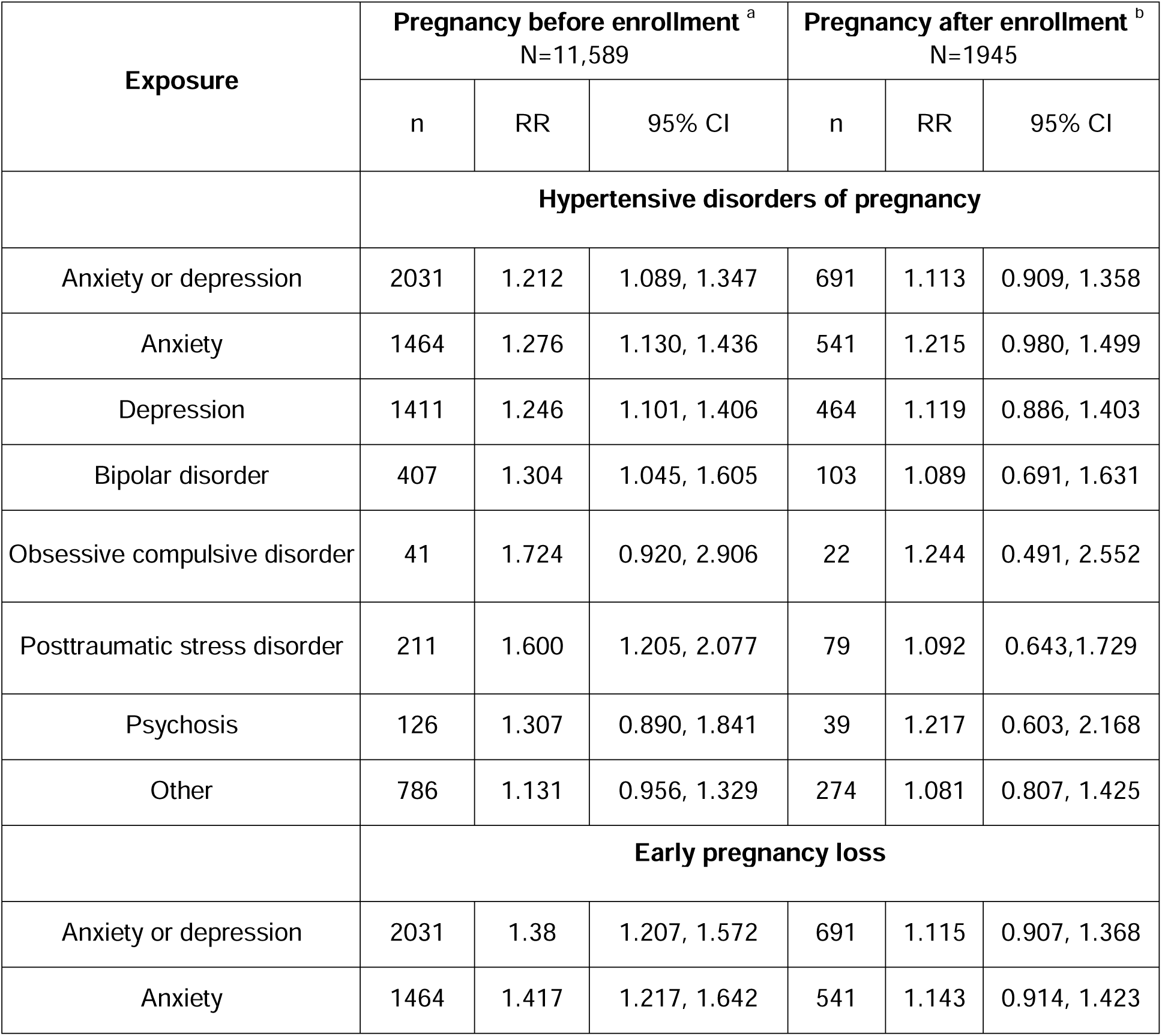
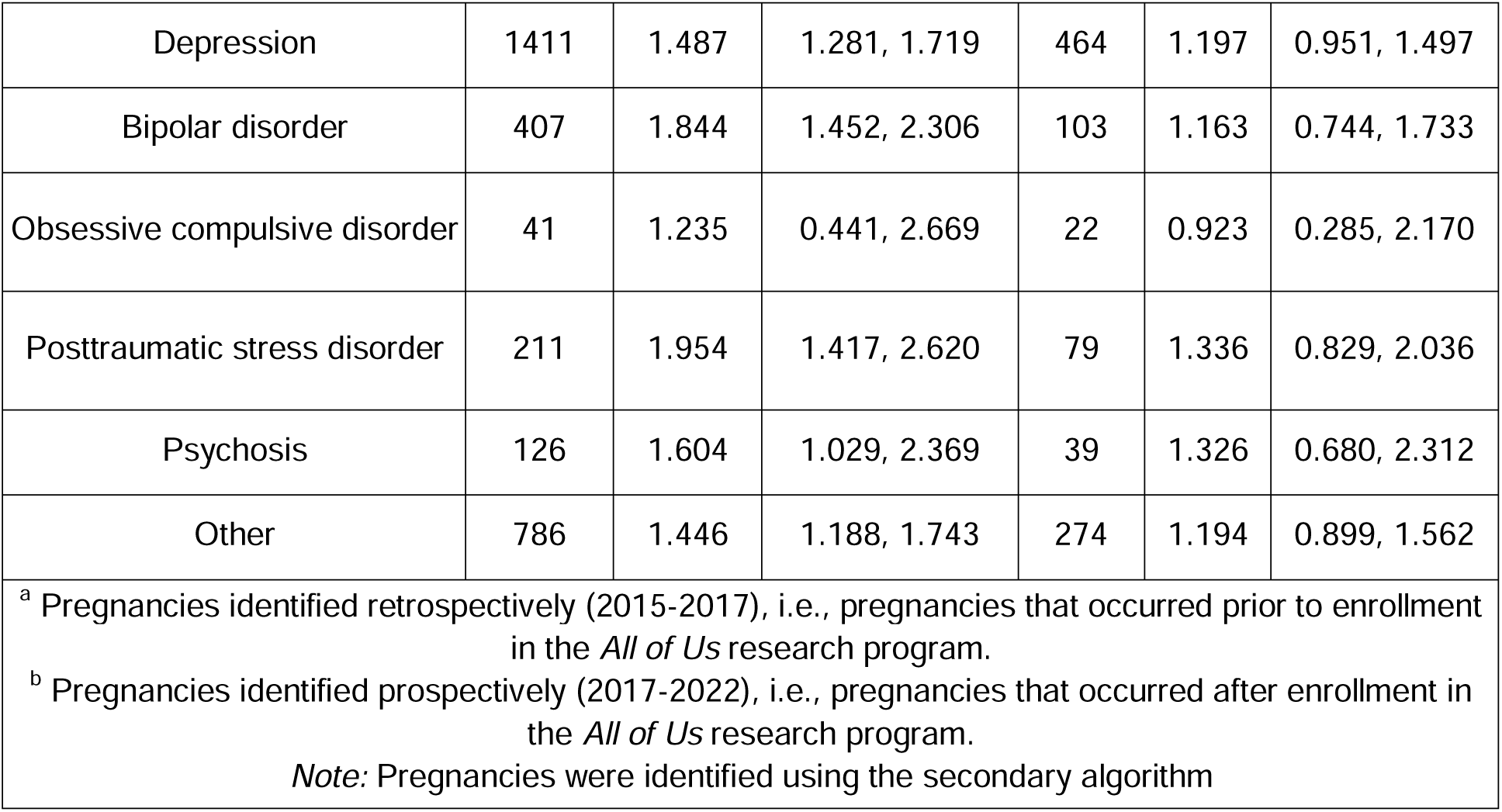
Association Between Diagnosed Mental Health Conditions and Hypertensive Disorders of Pregnancy or Early Pregnancy Loss, using the Secondary Algorithm to identify pregnancies, *All of Us* 2015-2022.

**Supplemental Figure 1.**
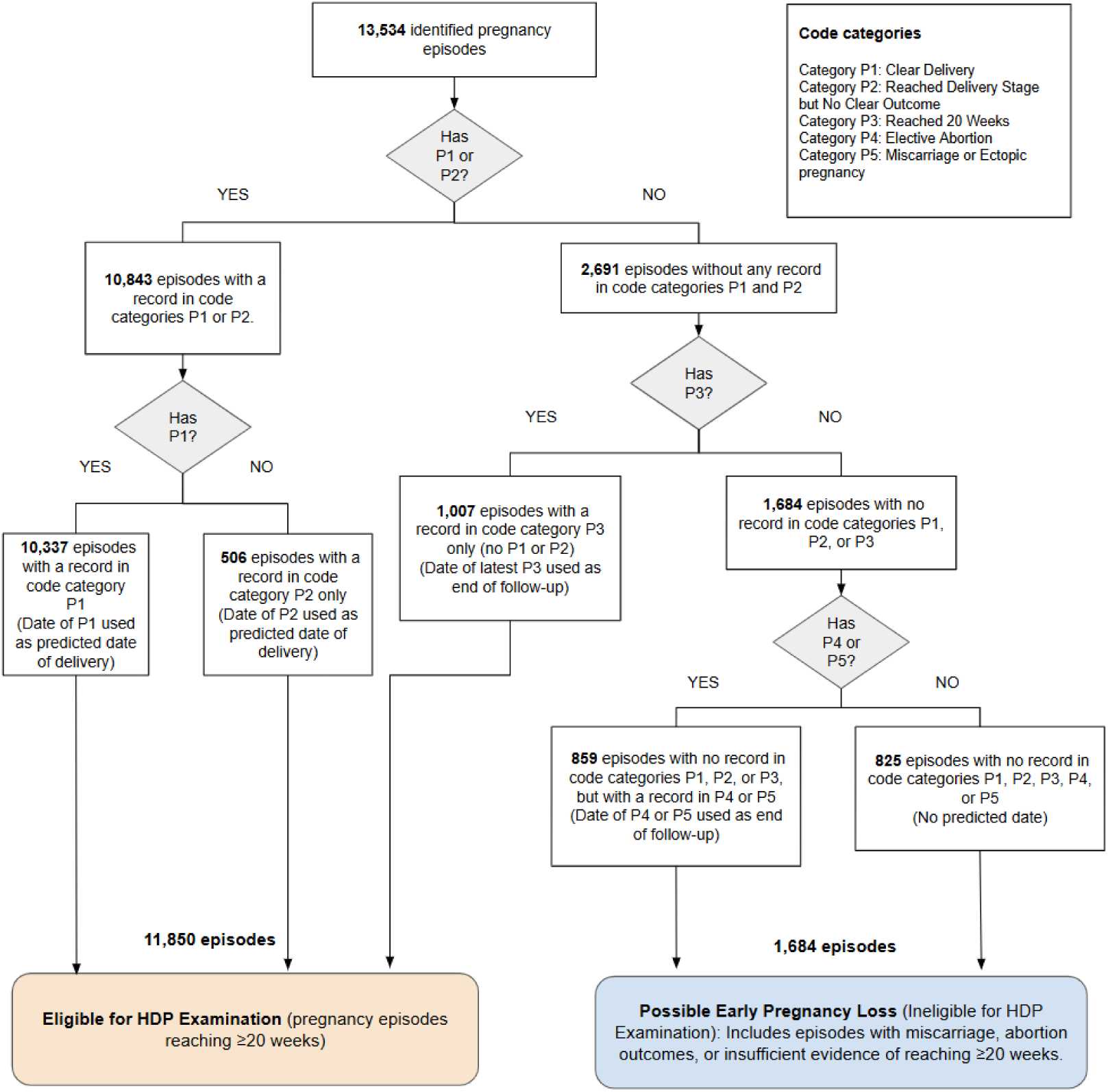
Decision Tree for Pregnancy Outcome Categorization in the Secondary Algorithm.

**Supplemental Figure 2.**
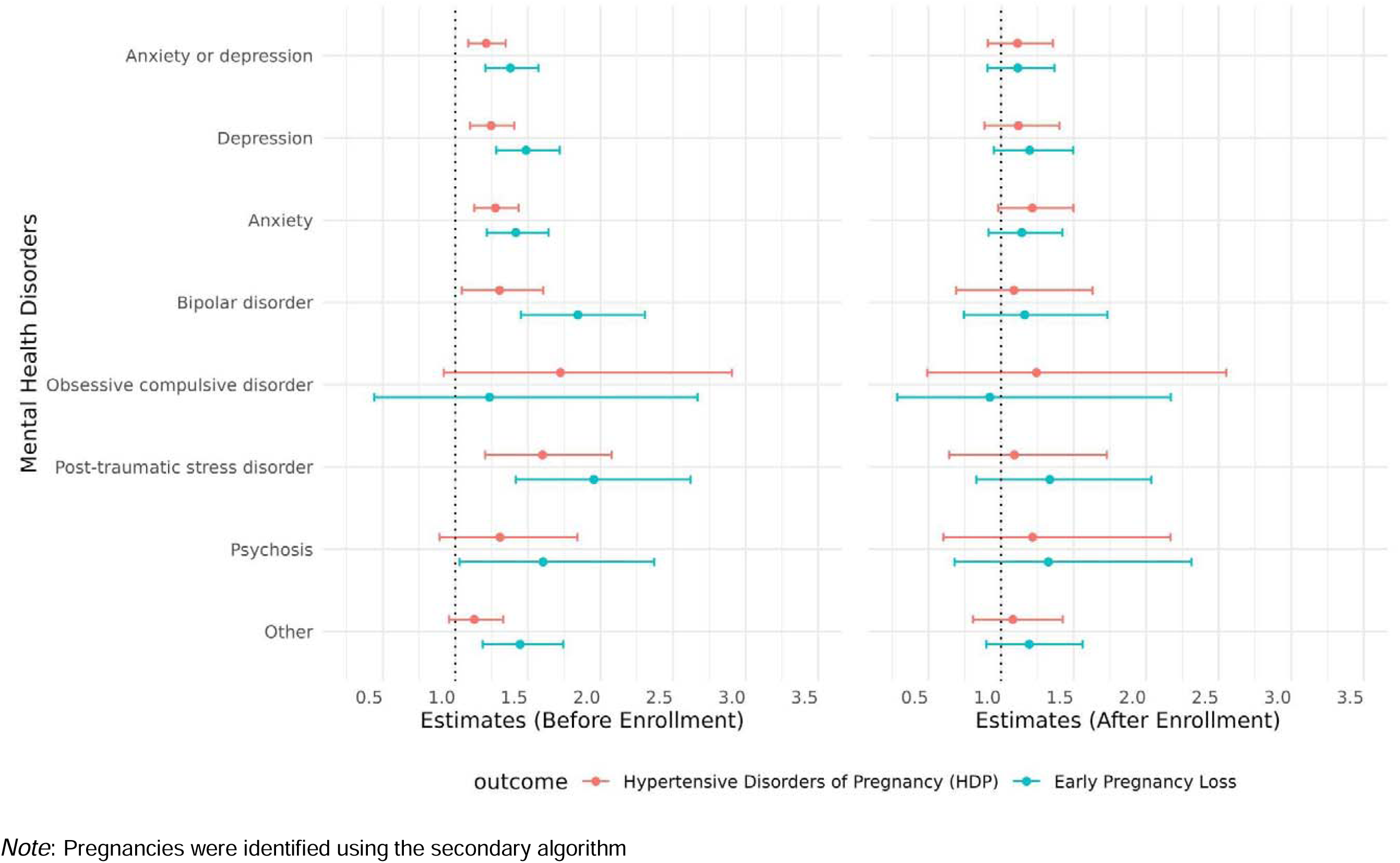
Association Between Diagnosed Mental Health Conditions and Hypertensive Disorders of Pregnancy or Early Pregnancy Loss, using the Secondary Algorithm to identify pregnancies, *All of Us* 2015-2022.

## References

1. Bruno AM, Allshouse AA, Metz TD, Theilen LH. Trends in hypertensive disorders of pregnancy in the U.S. from 1989 to 2018. Am J Obstet Gynecol. 2022;226(1):S495–S496. doi:10.1016/j.ajog.2021.11.819

2. Bellamy L, Casas JP, Hingorani AD, Williams DJ. Pre-eclampsia and risk of cardiovascular disease and cancer in later life: systematic review and meta-analysis. BMJ. 2007;335(7627):974. doi:10.1136/bmj.39335.385301.BE

3. Riise HKR, Sulo G, Tell GS, et al. Association Between Gestational Hypertension and Risk of Cardiovascular Disease Among 617 589 Norwegian Women. J Am Heart Assoc. 2018;7(10):e008337. doi:10.1161/JAHA.117.008337

4. Skjaerven R, Wilcox AJ, Klungsoyr K, et al. Cardiovascular mortality after pre-eclampsia in one child mothers: prospective, population based cohort study. BMJ. 2012;345(nov27 1):e7677-e7677. doi:10.1136/bmj.e7677

5. Langham J, Gurol-Urganci I, Muller P, et al. Obstetric and neonatal outcomes in pregnant women with and without a history of specialist mental health care: a national population-based cohort study using linked routinely collected data in England. Lancet Psychiatry. 2023;10(10):748–759. doi:10.1016/S2215-0366(23)00200-6

6. Elsaid N, Saied A, Kandil H, et al. Impact of stress and hypertension on the cerebrovasculature. Front Biosci-Landmark. 2021;26(12):1643. doi:10.52586/5057

7. Zhang S, Ding Z, Liu H, et al. Association between mental stress and gestational hypertension/preeclampsia: a meta-analysis. Obstet Gynecol Surv. 2013;68(12):825–834. doi:10.1097/OGX.0000000000000001

8. Liu MY, Na L, Li WA, Khan H. Association between psychosocial stress and hypertension: a systematic review and meta-analysis. Neurol Res. 2017;39(6):573–580. 10.1080/01616412.2017.1317904

9. O’Donnell MG, Stumpp L, Gallaher MJ, Powers RW. Pre-pregnancy stress induces maternal vascular dysfunction during pregnancy and postpartum. Reprod Sci. 2023;30(11):3197–3211. doi:10.1007/s43032-023-01248-2

10. Thombre Kulkarni M, Holzman C, Wasilevich E, Luo Z, Scheid J, Allswede M. Pregnancy hypertension and its associations with pre-pregnancy depression, anxiety, antidepressants, and anxiolytics. Pregnancy Hypertens. 2019;16:67–74. doi:10.1016/j.preghy.2019.03.003

11. Qiu C, Williams MA, Calderon-Margalit R, Cripe SM, Sorensen TK. Preeclampsia Risk in Relation to Maternal Mood and Anxiety Disorders Diagnosed Before or During Early Pregnancy. Am J Hypertens. 2009;22(4):397–402. doi:10.1038/ajh.2008.366

12. Thombre MK, Talge NM, Holzman C. Association Between Pre-Pregnancy Depression/Anxiety Symptoms and Hypertensive Disorders of Pregnancy. J Womens Health. 2015;24(3):228–236. doi:10.1089/jwh.2014.4902

13. Vollebregt K, van der Wal M, Wolf H, Vrijkotte T, Boer K, Bonsel G. Is psychosocial stress in first ongoing pregnancies associated with pre-eclampsia and gestational hypertension? Br J Obstet Gynaecol. 2008;115(5):607–615. doi:10.1111/j.1471-0528.2008.01665.x

14. Palmsten K, Setoguchi S, Margulis AV, Patrick AR, Hernández-Díaz S. Elevated Risk of Preeclampsia in Pregnant Women With Depression: Depression or Antidepressants? Am J Epidemiol. 2012;175(10):988–997. doi:10.1093/aje/kwr394

15. The “All of Us” Research Program. N Engl J Med. Published online 2019.

16. Data Snapshots. All of Us Research Hub. January 2025. https://www.researchallofus.org/data-tools/data-snapshots/

17. Smith LH, Wang W, Keefe-Oates B. Pregnancy episodes in All of Us: Harnessing multi-source data for pregnancy-related research.

18. Jones SE, Bradwell KR, Chan LE, et al. Who is pregnant? Defining real-world data-based pregnancy episodes in the National COVID Cohort Collaborative (N3C). JAMIA Open. 2023;6(3):ooad067. doi:10.1093/jamiaopen/ooad067

19. CMS. ICD-9-CM to and from ICD-10-CM and ICD-10-PCS Crosswalk or General Equivalence Mappings. NBER. https://www.nber.org/research/data/icd-9-cm-and-icd-10-cm-and-icd-10-pcs-crosswalk-or-general-equivalence-mappings

20. Perinatal Mental Health Conditions Patient Safety Bundle. AIM Alliance Innov Matern Health. Published online January 2024. https://saferbirth.org/wp-content/uploads/Perinatal-Mental-Health-Conditions-Patient-Safety-Bundle-2-4-1.pdf

21. Kircanski K, LeMoult J, Ordaz S, Gotlib IH. Investigating the nature of co-occurring depression and anxiety: Comparing diagnostic and dimensional research approaches. J Affect Disord. 2017;216:123–135. doi:10.1016/j.jad.2016.08.006

22. Labgold K, Stanhope KK, Joseph NT, Platner M, Jamieson DJ, Boulet SL. Validation of Hypertensive Disorders During Pregnancy: ICD-10 Codes in a High-burden Southeastern United States Hospital. Epidemiology. 2021;32(4):591–597. doi:10.1097/EDE.0000000000001343

23. Gunderson EP, Greenberg M, Nguyen-Huynh MN, et al. Early Pregnancy Blood Pressure Patterns Identify Risk of Hypertensive Disorders of Pregnancy Among Racial and Ethnic Groups. Hypertension. 2022;79(3):599–613. doi:10.1161/HYPERTENSIONAHA.121.18568

24. Solmi M, Radua J, Olivola M, et al. Age at onset of mental disorders worldwide: large-scale meta-analysis of 192 epidemiological studies. Mol Psychiatry. 2022;27(1):281–295. doi:10.1038/s41380-021-01161-7

25. RStudio Team (2020). RStudio: Integrated Development Environment for R. doi:RStudio, PBC, Boston

26. Caplan M, Kennan-Devlin LS, Freedman A, et al. Lifetime Psychosocial Stress Exposure Associated with Hypertensive Disorders of Pregnancy. Am J Perinatol. 2021;38(13):1412–1419. doi:10.1055/s-0040-1713368

27. Runkle JD, Risley K, Roy M, Sugg MM. Association Between Perinatal Mental Health and Pregnancy and Neonatal Complications: A Retrospective Birth Cohort Study. Womens Health Issues. 2023;33(3):289–299. doi:10.1016/j.whi.2022.12.001

28. Raina J, Elgbeili G, Montreuil T, et al. The effect of maternal hypertension and maternal mental illness on adverse neonatal outcomes: A mediation and moderation analysis in a U.S. cohort of 9 million pregnancies. J Affect Disord. 2023;326:11–17. doi:10.1016/j.jad.2023.01.052

29. Shay M, MacKinnon AL, Metcalfe A, et al. Depressed mood and anxiety as risk factors for hypertensive disorders of pregnancy: a systematic review and meta-analysis. Psychol Med. 2020;50(13):2128–2140. doi:10.1017/S0033291720003062

30. Serrano-Lomelin J, Smith GN, Davidge ST, et al. Associations of Diabetes, Mental Health, and Asthma with Hypertensive Disorders of Pregnancy: A Population-based Case-Control Study in Alberta, Canada. Pregnancy Hypertens. 2024;38:101172. doi:10.1016/j.preghy.2024.101172

31. Gold KJ, Dalton VK, Schwenk TL, Hayward RA. What causes pregnancy loss? Preexisting mental illness as an independent risk factor. Gen Hosp Psychiatry. 2007;29(3):207–213. doi:10.1016/j.genhosppsych.2007.02.002

